# Robust Disease Prognosis via Diagnostic Knowledge Preservation: A Sequential Learning Approach

**DOI:** 10.1101/2025.09.22.25336414

**Authors:** Haresh Rengaraj Rajamohan, Yanqi Xu, Weicheng Zhu, Richard Kijowski, Kyunghyun Cho, Krzysztof J Geras, Narges Razavian, Cem M. Deniz

## Abstract

Accurate disease prognosis is essential for patient care but is often hindered by the lack of long-term data. This study explores deep learning training strategies that utilize large, accessible diagnostic datasets to pretrain models aimed at predicting future disease progression in knee osteoarthritis (OA), Alzheimer’s disease (AD), and breast cancer (BC). While diagnostic pretraining improves prognostic task performance, naive fine-tuning for prognosis can cause ‘catastrophic forgetting,’ where the model’s original diagnostic accuracy degrades, a significant patient safety concern in real-world settings. To address this, we propose a sequential learning strategy with experience replay. We used cohorts with knee radiographs, brain MRIs, and digital mammograms to predict 4-year structural worsening in OA, 2-year cognitive decline in AD, and 5-year cancer diagnosis in BC. Our results showed that diagnostic pretraining on larger datasets improved prognosis model performance compared to standard baselines, boosting both the Area Under the Receiver Operating Characteristic curve (AUROC) (e.g., Knee OA external: 0.77 vs 0.747; Breast Cancer: 0.874 vs 0.848) and the Area Under the Precision-Recall Curve (AUPRC) (e.g., Alzheimer’s Disease: 0.752 vs 0.683). Additionally, a sequential learning approach with experience replay achieved prognostic performance comparable to dedicated single-task models (e.g., Breast Cancer AUROC 0.876 vs 0.874) while also preserving diagnostic ability. This method maintained high diagnostic accuracy (e.g., Breast Cancer Balanced Accuracy 50.4% vs 50.9% for a dedicated diagnostic model), unlike simpler multitask methods prone to catastrophic forgetting (e.g., 37.7%). Our findings show that leveraging large diagnostic datasets is a reliable and data-efficient way to enhance prognostic models while maintaining essential diagnostic skills.

**Author Summary:** In our research, we addressed a common problem in medical AI: how to accurately predict the future course of a disease when long-term patient data is rare. We focused on knee osteoarthritis, Alzheimer’s disease, and breast cancer. We found that we could significantly improve a model’s ability to predict disease progression by first training it on a much larger, more common type of data - diagnostic images used to assess a patient’s current disease state. We then developed a specialized training method that allows a single AI model to perform both diagnosis and prognosis tasks effectively. A key challenge is that models often “forget” their original diagnostic skills when they learn a new prognostic task. In a clinical setting, this poses a safety risk, as it could lead to missed diagnoses. We utilize experience replay to overcome this by continually refreshing the model’s diagnostic knowledge. This creates a more robust and efficient model that mirrors a clinician’s workflow, offering the potential to improve patient care with limited amount of hard-to-get longitudinal data.

## 1 Introduction

Progressive diseases pose significant challenges to healthcare systems worldwide. Their gradual onset and potential for irreversible damage make early intervention, guided by accurate prognosis, crucial for optimizing patient outcomes, reducing healthcare costs, and improving quality of life [1, 2, 3]. Key examples include knee osteoarthritis (OA), Alzheimer’s disease (AD), and breast cancer (BC) - conditions representing diverse physiological systems but sharing the critical need for early identification and reliable progression prediction [1, 3, 4, 5].

Deep learning (DL) [6] offers powerful tools for automating medical image analysis and has shown considerable promise for both diagnosis and prognosis tasks across various diseases. Studies have demonstrated DL’s effectiveness in diagnosing OA severity [7], detecting AD from neuroimaging [8, 9, 10, 11], and screening for BC [12, 13]. Similarly, DL models have been developed to predict disease progression, such as total knee replacement risk in OA [14, 15, 16, 17, 18], progression to AD [19, 20], and long-term BC outcomes [21, 22].

However, developing robust prognosis models faces a significant hurdle: the scarcity of longitudinal data required to track disease progression over time. This contrasts with diagnosis, where larger cross-sectional datasets are often more readily available. To mitigate this data scarcity, previous works have explored multitask learning (MTL) as additional regularization, where a single model is trained to perform several related tasks simultaneously, aiming to improve generalization and performance through shared representations [13, 14, 16, 21, 22]. For instance, MTL combining diagnosis and prognosis has shown benefits in OA [14, 16], and pretraining on BI-RADS has been shown to improve current cancer diagnosis performance [13]. Yet these MTL studies typically rely on relatively small prognosis datasets and do not fully capitalize on the abundance of diagnostic data. We hypothesize that sequential learning - diagnosis pretraining on large, unbiased datasets followed by prognostic training, achieves a favorable balance of prognostic and diagnostic performance.

A key challenge arises, however, when building a single model that learns tasks sequentially. When a model pretrained on a data-rich task (e.g., diagnosis) is subsequently fine-tuned on a new, data-scarce task (e.g., prognosis), it often suffers from catastrophic forgetting, a significant degradation of its ability to perform the original task [29]. In the context of healthcare, this phenomenon is not merely a technical limitation but a critical issue for real-world model maintenance and patient safety. A prognostic model that compromises diagnostic accuracy could lead to missed disease during routine follow-up, undermining its clinical utility and trust. A primary strategy to combat this phenomenon is experience replay (or rehearsal), where the model is periodically retrained on stored examples from the original task while learning the new one, thereby refreshing and preserving its previously acquired knowledge [30].

We demonstrate that pretraining models on larger diagnostic datasets significantly improves prognostic performance, generalization to external cohorts, and discrimination within patient subgroups compared to standard initialization techniques across Knee OA, Alzheimer’s disease and breast cancer. We also identify that a sequential multitask learning approach with experience replay yields the best overall results, matching the prognostic performance of dedicated single-task models while effectively preserving diagnostic capabilities and outperforming simpler joint training strategies.

## 2 Materials and Methods

### 2.1 Ethics Statement

This retrospective study was conducted using data acquired under protocols approved by the relevant Institutional Review Boards (IRBs). The Osteoarthritis Initiative (OAI), Multicenter Osteoarthritis Study (MOST), and Alzheimer’s Disease Neuroimaging Initiative (ADNI) are publicly available research datasets for which all participants provided informed consent at the time of original data collection. The breast cancer dataset was sourced from New York University Langone Health under an IRB-approved protocol with a waiver of informed consent, in compliance with the Health Insurance Portability and Accountability Act (HIPAA).

### 2.2 Prediction Task Definitions

We investigate the following clinical measures for knee OA, Alzheimer’s Disease and Breast Cancer:

#### 2.2.1 Knee Osteoarthritis (OA)

**Diagnosis:** Classification based on Kellgren-Lawrence Grade (KLG) [26] - A severity scale (0-4) assigned by radiologists based on radiographic features including joint space narrowing, bone spurs, and sclerosis.

**Prognosis:** Prediction of disease worsening over a 4-year horizon [27], defined as:

- Structural Incidence for early-stage OA (KLG 0-1): Progression to KLG ≥ 2 or undergoing a Total Knee Replacement (TKR).
- Structural Progression for radiographic OA (KLG ≥ 2): An increase in KLG or undergoing a TKR.

#### 2.2.2 Alzheimer’s Disease (AD)

**Diagnosis:** Classification of cognitive status using structural brain MRI, based on the diagnostic criteria established by the ADNI study protocol [25]:

- Cognitive Normal (CN): Participants with no significant impairment in cognitive functions or activities of daily living.
- Mild Cognitive Impairment (MCI): Participants with a subjective memory concern, objective memory loss, and preserved general cognition and functional performance.
- Alzheimer’s Disease (AD): Participants meeting the NINCDS/ADRDA criteria for probable AD.

**Prognosis:** Prediction of cognitive decline over a 2-year horizon:

- CN → MCI: Onset of cognitive impairment.
- MCI → AD or CN → AD: Progression to confirmed disease.

#### 2.2.3 Breast Cancer (BC)

**BI-RADS (Diagnosis):** Classification system for breast imaging findings:

- Grade 0: Incomplete assessment; additional imaging evaluation is needed.
- Grade 1: Negative; no abnormalities found.
- Grade 2: Benign finding; no malignant characteristics present.

**Prognosis:** Prediction of a patient having a confirmed breast cancer diagnosis within 5 years of the scan date.

### 2.3 Datasets and Cohorts

#### 2.3.1 Knee Osteoarthritis

Data were sourced from the **Osteoarthritis Initiative (OAI)** [23] for model development and the **Multicenter Osteoarthritis Study (MOST)** [24] for external validation. The OAI is a long-term, multicenter observational study of 4,796 participants focused on knee OA biomarkers. We utilized bilateral posterior-anterior fixed flexion knee radiographs.

- **Diagnosis Cohort (OAI):** Comprised 47,041 radiographs from 4,508 subjects with KLG assessments.
- **Prognosis Cohort (OAI):** To predict structural worsening, a 4 year follow up from the baseline visit was used. This timeframe was selected to balance observing meaningful change and maximizing cohort size, as longer horizons led to significant participant drop-off. Further, knees with KLG 4 or a TKR at baseline were excluded. Two distinct prognosis tasks and their corresponding cohorts were defined based on baseline disease severity:

- **Structural Incidence:** This task focused on predicting the onset of radiographic OA in knees with baseline KLG 0-1. The cohort included 420 patients who progressed (defined as reaching KLG ≥ 2 or undergoing a TKR) and 2,439 non-progressing controls.
- **Structural Progression:** This task focused on predicting the worsening of established OA in knees with baseline KLG 2-3. The cohort included 571 patients who progressed (defined as an increase in KLG or undergoing a TKR) and 1,677 non-progressing controls.
- **MOST Validation Cohort:** Corresponding cohorts were created from the MOST dataset using a 5-year follow-up due to data availability. The incidence cohort included 505 progressors and 1,395 controls. The progression cohort included 562 progressors and 617 controls.

#### 2.3.2 Alzheimer’s Disease

Data were obtained from the **Alzheimer’s Disease Neuroimaging Initiative** (ADNI) database [25], a longitudinal multicenter study aimed at validating biomarkers for AD progression. We utilized T1-weighted brain MRI scans.

- **Diagnosis Cohort:** Consisted of 2,723 scans from 662 patients classified as Cognitive Normal (CN), Mild Cognitive Impairment (MCI), or Alzheimer’s Disease (AD).
- **Prognosis Cohort:** To predict cognitive decline within a 24-month follow-up, 914 scans from 365 unique patients were used. Progression was defined as a change from CN to MCI/AD or MCI to AD. To augment the dataset size, MRI scans from both baseline and available intermediate follow-up visits were utilized as distinct input time points; however, scans from patients already classified as AD were excluded. To prevent data leakage and ensure the model learned generalizable prognostic features rather than patient-specific anatomy, all scans from a single patient were strictly kept within the same data partition (training, validation, or test) during all experiments. The cohort included 148 progressing patients (contributing 343 scans) and 217 stable controls (contributing 571 scans).

#### 2.3.3 Breast Cancer

We utilized a curated dataset of digital screening mammograms from NYU Langone Health [28], enhanced with longitudinal follow-up cancer labels up to five years post-imaging.

- **Diagnosis Cohort:** Contained 56,733 examinations from 33,681 patients with radiologist-assigned BI-RADS grades 0, 1, or 2.
- **Prognosis Cohort:** To facilitate robust model development, a balanced cohort was constructed. This included 3,000 examinations from 2,319 patients who were diagnosed with breast cancer within 5 years (but more than 130 days after the index scan) and 3,000 control examinations from 2,837 patients who remained cancer-free for the 5-year follow-up period. The mean age for patients contributing case examinations was 61.5 ± 11.4 years, while the mean age for controls was 56.4 ± 10.8 years.

To prevent data leakage, strict patient-level splits were used to ensure disjoint sets of individuals across all training, validation, and test sets, and no information from future visits was used as input features. Detailed demographic and clinical characteristics for all patient cohorts are provided in S1-S4 Tables.

### 2.4 Approach

Our work investigates two key approaches: utilizing diagnosis tasks for model pretraining and proposing an effective multitask learning approach.

#### 2.4.1 Diagnosis Pretraining

We compare diagnosis-based initialization against modality-specific baselines (Fig. 1). We use modality-appropriate initializations: OA is 2D and moderate-resolution, where ImageNet pretraining is standard; AD uses 3D brain MRI (no ImageNet analogue), and BC requires very high-resolution mammography where running large ImageNet backbones at native resolution is not tractable, so we use random initialization similar to previous works [8, 12]. Concretely, prognosis models are initialized from either (i) diagnosis-pretrained weights learned on the corresponding diagnostic task, (ii) **ImageNet weights** (OA only), or (iii) **random weights** (AD, BC).

**Fig 1:**
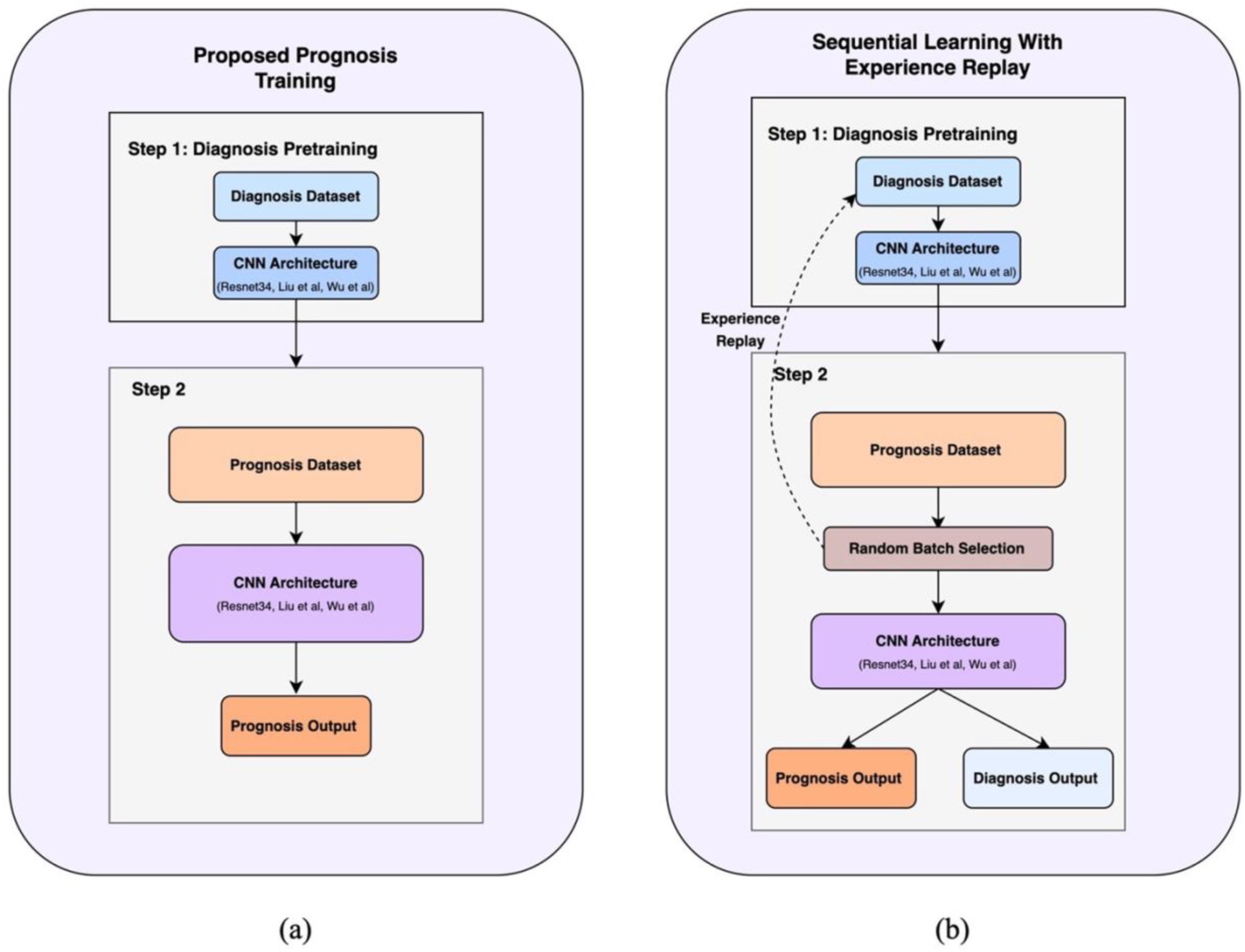
Illustrates the proposed approaches (a) single task prognosis with diagnostic pretraining and (b) Sequential learning with experience replay

#### 2.4.2 Sequential Learning with Experience Replay

To mitigate catastrophic forgetting [29, 30] in multitask settings, we used a two-phase approach (Fig 1).

1. **Phase 1 (Diagnosis Pretraining):** A model is trained on the large diagnosis cohort.
2. **Phase 2 (Prognosis training with Diagnosis Replay):** The model is fine-tuned using a multitask objective. At each step, a batch is randomly sampled with equal probability from either the prognosis cohort (to update on the prognosis task) or the diagnosis cohort (to “replay” and refresh diagnosis knowledge). This allows the model to learn the new task while preserving its original capabilities.

#### 2.4.3 Comparative Multitask Approaches

For comparison, we evaluated three alternative strategies:

- **Single-cohort MT:** An approach similar to that used by Tiulpin et al. [14] and Leung et al.[16], where the model is trained on both diagnosis and prognosis tasks using only the data available in the smaller prognosis cohort.
- **Diagnosis-pretrained MT:** Initializes with diagnostic pretraining but then fine-tunes solely on the prognosis cohort’s diagnosis and prognosis labels, without experience replay
- **Concurrent MT:** Trains simultaneously on both diagnosis and prognosis cohorts from a baseline initialization, without any pretraining.

### 2.5 Experimental Setup

#### 2.5.1 Model Architectures and Preprocessing

- **OA:** Input radiographs were normalized, and knee joints were extracted prior to analysis. ResNet-34 with attention [31, 32] architecture was used on extracted knee joints.
- **AD:** Input MRI scans underwent standard bias correction and spatial normalization. A custom 3D-CNN [8] was used on processed T1 MRIs.
- **BC:** The four standard mammographic views were individually processed and cropped to a uniform resolution. A custom multi-view CNN [12] was used on the processed mammogram views.

#### 2.5.2 Training Protocol

All models were trained using the Adam optimizer [33]. A cosine annealing learning rate scheduler was used to ensure stable convergence. For the highly imbalanced breast cancer BI-RADS task, a weighted cross-entropy loss was employed to give appropriate importance to rare classes. Condition-specific data augmentation techniques (including random cropping, flips, and rotations) were applied during training to enhance model robustness.

For tasks with smaller sample sizes (all prognosis cohorts and the AD diagnosis cohort), we employed a 5-fold cross-validation scheme with strict, non-overlapping patient-level splits. For each of the 5 iterations, one fold was designated as the held-out test set. Of the remaining four folds, three were used for training the model, and one was used as the validation set for hyperparameter tuning and model checkpointing via early stopping. This process was repeated five times, ensuring that every patient served as part of a test set exactly once. The final reported performance metrics are the mean and standard deviation across the results from the five test folds.

For multitask learning with experience replay, tasks were sampled with equal probability (p=0.5). Crucially, to prioritize the more challenging prognostic task, model checkpoints were saved based on the epoch achieving the highest prognosis AUROC on the validation set, rather than on the total loss

For each disease, the diagnosis and prognosis cohorts were drawn from the same underlying patient populations (OAI, ADNI, and NYU dataset). As such, patients in the longitudinal prognosis cohort were also represented in the larger cross-sectional diagnosis cohort. To prevent any data leakage between the pretraining and fine-tuning stages, a strict patient-level splitting procedure was enforced. For each of the 5 cross-validation folds of the prognosis task, the patient IDs assigned to the test set were identified first. These testset patients were then completely excluded from the training set of the large diagnosis model used for pretraining. This ‘test set holdout’ strategy ensures that our prognosis models were always evaluated on patients that the entire training pipeline, including the pretraining stage, had never seen, providing an unbiased estimate of performance.

#### 2.5.3 Evaluation and Analysis

This study was prepared and reported in accordance with the TRIPOD (Transparent Reporting of a multivariable prediction model for Individual Prognosis or Diagnosis) guideline wherever applicable. The pre-specified primary endpoint for all prognostic tasks was the Area Under the Receiver Operating Characteristic curve (AUROC), which measures the model’s overall ability to discriminate between patients who experience disease progression and those who do not.

Secondary endpoints for prognosis included the Area Under the Precision-Recall Curve (AUPRC), which provides additional insight into model performance, especially in cohorts with class imbalance. Sensitivity and specificity were computed additionally for breast cancer.

For the threshold-dependent metrics of sensitivity and specificity, the following procedure was used for each of the 5 cross-validation folds: first, an optimal decision threshold was determined by maximizing the Youden Index [34] on the validation set for that fold. This fold-specific threshold was then applied to the predictions on the corresponding test set to calculate sensitivity and specificity. The final reported metrics are the mean and standard deviation of the five resulting sensitivity and specificity values.

For diagnosis, we report balanced accuracy for the five-class knee OA task, following [16] to mitigate class imbalance. For the multi-class AD and BC tasks, we additionally report macro-AUROC and micro-AUROC, consistent with prior work. Macro-AUROC is the unweighted mean of one-vs-rest AUROCs across classes, giving each class equal weight. Micro-AUROC pools predictions and labels across all classes to compute a single AUROC, effectively weighting classes by their prevalence. Balanced accuracy is the mean of per-class recall (sensitivity).

To formally compare the primary approaches, ROC curves of ensembled cross-validation predictions were compared using the DeLong test [35]. For every patient in the test set, the predictive scores (probabilities) from the five respective cross-validation models were averaged. This resulted in a single set of ensembled predictions for the entire test cohort, from which a single, more stable ROC curve was generated for the statistical comparison. This practice of averaging predictions is a standard ensembling technique to reduce variance and provide a more robust estimate of model performance.

Stratified subgroup analyses were performed on pre-defined clinical subgroups: structural incidence (KL 0-1) vs. progression (KL 2-3) for Knee OA, and cognitive transition groups (e.g., CN vs. MCI) for Alzheimer’s Disease. These subgroup analyses were conducted to verify that the models were learning genuine prognostic signals rather than simply using baseline disease severity as a proxy for progression risk. This analysis was not powered for formal statistical tests of subgroup interaction. Statistical significance testing was focused on our primary hypothesis comparing the baseline and diagnosis-pretrained models.

## 3 Results

The experimental results are presented in two main parts. The first subsection evaluates the impact of initializing prognosis models with weights pretrained on large diagnostic datasets, comparing their performance against standard baselines for single-task prognosis. The second subsection compares four multitask learning strategies on both prognosis and diagnosis.

### 3.1 Impact of Diagnosis Pretraining

Initializing prognosis models with weights pretrained on diagnostic tasks with full diagnosis cohorts, yielded improvements over baseline initializations (ImageNet for Knee OA, random for AD/BC), as detailed in Table 1.

**Table 1.**
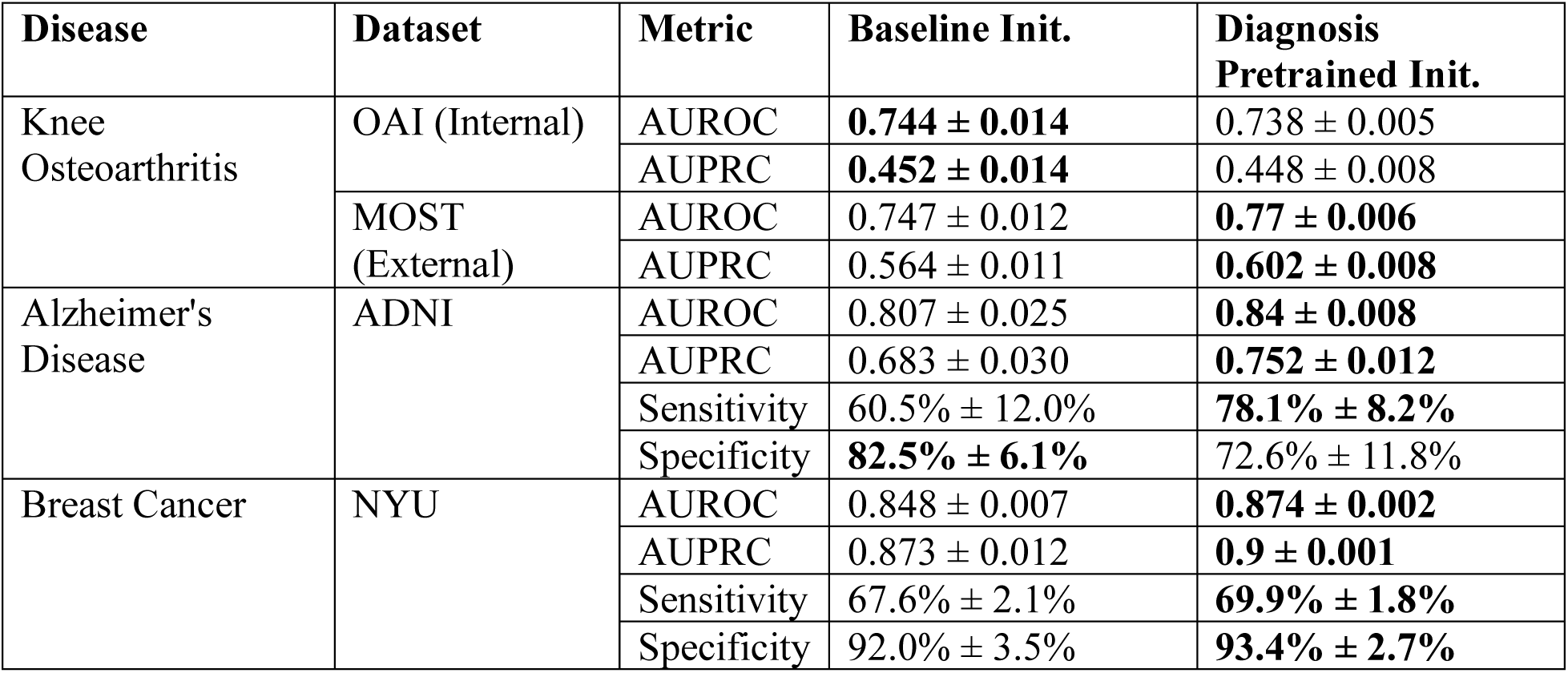
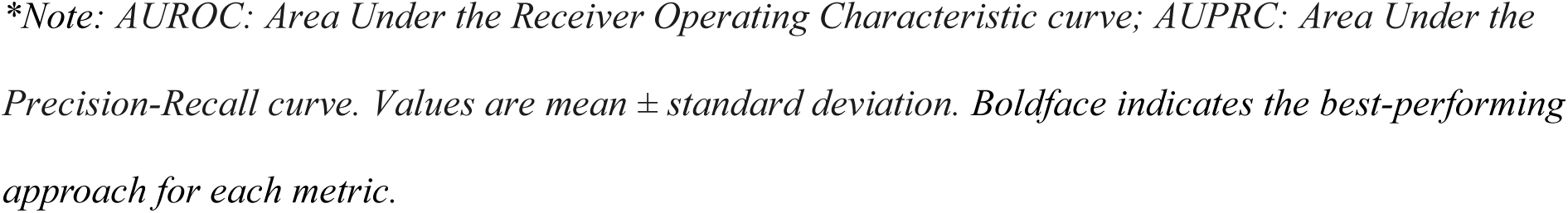
Comparison of prognosis prediction performance using models with baseline initialization versus diagnostic pretraining across three diseases. Knee OA models used ImageNet pretraining as baseline; AD and Breast Cancer models used random initialization as baseline.

| Disease | Dataset | Metric | Baseline Init. | Diagnosis Pretrained Init. |
| --- | --- | --- | --- | --- |
| Knee Osteoarthritis | OAI (Internal) | AUROC | **$0.744 \pm 0.014$** | $0.738 \pm 0.005$ |
| | | AUPRC | **$0.452 \pm 0.014$** | $0.448 \pm 0.008$ |
| | MOST (External) | AUROC | $0.747 \pm 0.012$ | **$0.77 \pm 0.006$** |
| | | AUPRC | $0.564 \pm 0.011$ | **$0.602 \pm 0.008$** |
| Alzheimer's Disease | ADNI | AUROC | $0.807 \pm 0.025$ | **$0.84 \pm 0.008$** |
| | | AUPRC | $0.683 \pm 0.030$ | **$0.752 \pm 0.012$** |
| | | Sensitivity | $60.5\% \pm 12.0\%$ | **$78.1\% \pm 8.2\%$** |
| | | Specificity | **$82.5\% \pm 6.1\%$** | $72.6\% \pm 11.8\%$ |
| Breast Cancer | NYU | AUROC | $0.848 \pm 0.007$ | **$0.874 \pm 0.002$** |
| | | AUPRC | $0.873 \pm 0.012$ | **$0.9 \pm 0.001$** |
| | | Sensitivity | $67.6\% \pm 2.1\%$ | **$69.9\% \pm 1.8\%$** |
| | | Specificity | $92.0\% \pm 3.5\%$ | **$93.4\% \pm 2.7\%$** |

*\*Note: AUROC: Area Under the Receiver Operating Characteristic curve; AUPRC: Area Under the Precision-Recall curve. Values are mean ± standard deviation. Boldface indicates the best-performing approach for each metric.*

For Knee Osteoarthritis, while performance on the internal OAI dataset was comparable, diagnosis-pretrained models demonstrated superior generalization on the external MOST dataset (e.g., AUROC 0.77 vs 0.747) and exhibited lower variance, indicating more stable performance. This improved generalization on the external MOST dataset was statistically significant when comparing model ensembles via DeLong’s test (AUROC 0.781 for diagnosis-pretrained vs. 0.768 for ImageNet-pretrained, p=0.006, n=4251). This performance benefit on the external MOST dataset was consistent across both early-stage (Incidence) and established (Progression) disease subgroups (Fig 2a).

**Fig 2.**
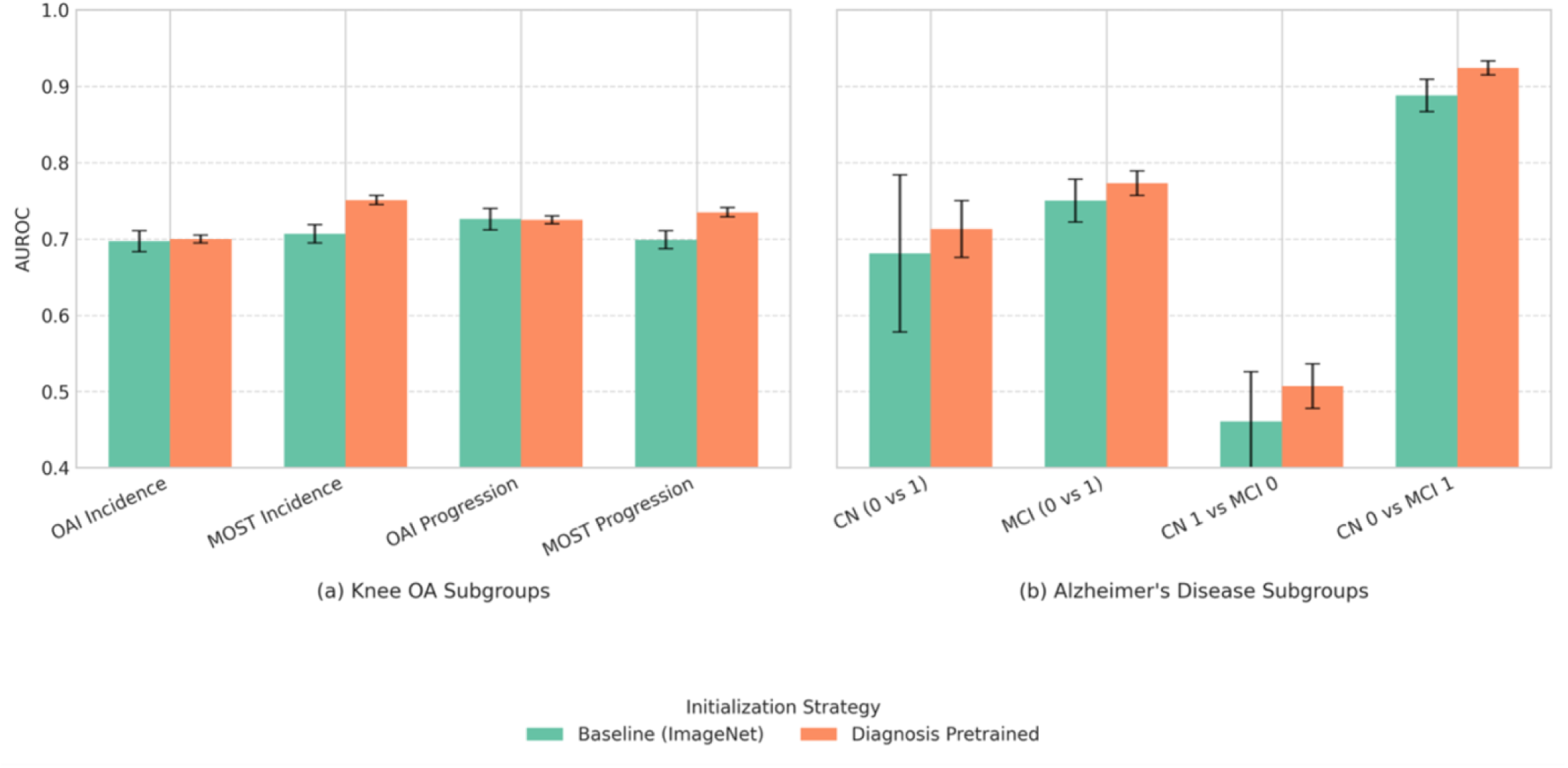
Comparison of baseline initialization versus diagnostic pretraining for prognosis prediction performance (AUROC) within specific disease subgroups. (a) Knee OA subgroups include Incidence (progression from KL 0-1) and Progression (worsening from KL 2-3) cohorts evaluated on the internal OAI and external MOST datasets. (b) Alzheimer’s Disease subgroups represent different cognitive transitions within the ADNI dataset (CN: Cognitively Normal, MCI: Mild Cognitive Impairment; 0: Stable status over 2 years, 1: Progressing status over 2 years). Diagnostic pretraining generally improves or maintains performance with better stability, showing particular benefit in the external MOST and all AD sub cohorts. Full numerical results are available in S5 and S6 Tables.

For Alzheimer’s Disease, diagnostic pretraining significantly enhanced AD progression prediction across all metrics, notably increasing AUROC (0.84 vs 0.807) and AUPRC (0.752 vs 0.683). Benefits were observed across different cognitive subgroups (Fig 2b). However, the difference in overall AUROC between model ensembles on the whole test set was not statistically significant via DeLong’s test (0.846 for diagnosis-pretrained vs. 0.823 for random initialized, p=0.137, n=236).

For Breast Cancer, diagnostic pretraining substantially improved 5-year prognosis prediction (AUROC 0.874 vs 0.848). This boost extended to precision-recall (AUPRC 0.9 vs 0.873) and clinical utility metrics (sensitivity/specificity), alongside improved stability (lower std. dev.). This performance increase was statistically significant when comparing model ensembles (AUROC 0.876 for diagnostic-pretrained vs. 0.854 for random initialized, p<0.001, n=1735).

Across all three diseases, these findings reinforce that leveraging diagnostic knowledge provides a valuable initialization for prognostic tasks, likely by helping the model learn representations of relevant anatomical and pathological features that precede clinically evident progression.

### 3.2 Multitask Learning Performance

We further compared four multitask learning strategies, evaluating performance on both the prognosis task (Table 2) and the diagnosis task (Table 3).

**Table 2.** Prognosis prediction performance across different multitask learning approaches. ‘Ref (Diag. Pret)’ refers to the single-task prognosis model initialized with diagnostic pretraining (from Table 1). ‘Single-cohort MT’ uses only prognosis cohort data for multitask training. ‘Concurrent MT’ trains on diagnosis and prognosis cohorts simultaneously from baseline initialization. ‘Diag-pret MT’ performs diagnostic pretraining then trains multitask on the prognosis cohort. ‘Seq Learn w/ Replay’ initializes with diagnosis pretraining, trains multitask on prognosis cohort and uses experience replay with the diagnosis cohort.

| Disease | Dataset | Metric | Single-task prognosis (diagnosis-pretrained init.) | Single-cohort MT | Concurrent MT | Diag-pret MT | Seq Learn w/ Replay |
| --- | --- | --- | --- | --- | --- | --- | --- |
| Knee Osteoarthritis | OAI (Internal) | AUROC | 0.738 ± 0.005 | 0.739 ± 0.017 | 0.743 ± 0.011 | 0.748 ± 0.005 | **0.75 ± 0.006** |
| | | AUPRC | 0.448 ± 0.008 | 0.444 ± 0.015 | 0.438 ± 0.011 | **0.458 ± 0.005** | 0.456 ± 0.006 |
| | MOST (External) | AUROC | **0.770 ± 0.006** | 0.756 ± 0.027 | 0.765 ± 0.005 | 0.763 ± 0.007 | 0.764 ± 0.007 |
| | | AUPRC | **0.602 ± 0.008** | 0.564 ± 0.020 | 0.589 ± 0.004 | 0.599 ± 0.008 | 0.594 ± 0.014 |
| Alzheimer's Disease | ADNI | AUROC | 0.840 ± 0.008 | 0.798 ± 0.014 | 0.828 ± 0.016 | 0.848 ± 0.008 | **0.851 ± 0.009** |
| | | AUPRC | 0.752 ± 0.012 | 0.653 ± 0.027 | 0.716 ± 0.021 | **0.754 ± 0.011** | **0.754 ± 0.005** |
| Breast Cancer | NYU | AUROC | 0.874 ± 0.002 | 0.864 ± 0.002 | 0.860 ± 0.007 | **0.878 ± 0.003** | 0.876 ± 0.003 |
| | | AUPRC | 0.900 ± 0.001 | 0.894 ± 0.002 | 0.889 ± 0.008 | **0.903 ± 0.003** | **0.903 ± 0.002** |
| | | Sensitivity | **69.9% ± 1.8%** | 66.1% ± 1.6% | 67.4% ± 2.6% | 69.0% ± 2.5% | 67.7% ± 1.7% |
| | | Specificity | 93.4% ± 2.7% | 97.1% ± 1.0% | 94.6% ± 4.3% | 95.5% ± 1.8% | **97.4% ± 1.2%** |

*\*Note: Values are mean ± standard deviation. Boldface indicates the best-performing approach for each metric.*

**Table 3.** Diagnosis task performance across different multitask learning approaches. ‘Reference’ refers to a model trained only on the diagnosis task. Performance is measured by Balanced Accuracy (Bal Acc) for Knee OA; and Macro AUROC, Micro AUROC, and Bal Acc for Alzheimer’s Disease (AD) and Breast Cancer (BC). Other approach definitions are as in Table 2.

| Disease Condition | Dataset | Metric | Reference | Single-cohort MT | Concurrent MT | Diag-pret MT | Seq Learn w/ Replay |
| --- | --- | --- | --- | --- | --- | --- | --- |
| Knee Osteoarthritis | OAI (Internal) | Bal Acc (%) | **75.1%** | 51.9% ± 1.3% | 73.4% ± 2.4% | 56.6% ± 0.7% | **75.1% ± 1.1%** |
| | MOST (External) | Bal Acc (%) | 65.6% | 47.0% ± 3.0% | **67.3% ± 1.2%** | 48.8% ± 1.7% | **67.5% ± 2.6%** |
| Alzheimer's Disease | ADNI | Macro AUROC | **0.752 ± 0.012** | 0.530 ± 0.011 | 0.741 ± 0.013 | 0.532 ± 0.026 | **0.758 ± 0.005** |
| | | Micro AUROC | 0.76 ± 0.016 | 0.581 ± 0.007 | 0.746 ± 0.007 | 0.618 ± 0.015 | **0.763 ± 0.011** |
| | | Bal Acc (%) | **56.3% ± 4.2%** | 35.3% ± 3.2% | 52.7% ± 3.0% | 38.1% ± 2.9% | 53.0% ± 2.8% |
| Breast Cancer | NYU | Macro AUROC | **0.7** | 0.577 ± 0.007 | 0.571 ± 0.007 | 0.657 ± 0.004 | **0.697 ± 0.002** |
| | | Micro AUROC | **0.724** | 0.687 ± 0.002 | 0.655 ± 0.013 | 0.726 ± 0.012 | **0.735 ± 0.012** |
| | | Bal Acc (%) | **50.9%** | 37.7% ± 1.2% | 41.3% ± 0.4% | 45.2% ± 0.4% | **50.4% ± 0.6%** |

*\*Note: Values are reported as mean ± standard deviation where available from cross-validation, otherwise as single run results. Boldface indicates the best-performing approach for each metric.*

For Knee Osteoarthritis, Sequential Learning with Experience Replay and Diagnosis-pretrained MT demonstrated the strongest prognostic performance (e.g., OAI AUROC 0.75, MOST AUROC 0.764), matching the dedicated single-task reference model. On the diagnosis task, Sequential Learning with Experience Replay maintained robust performance (OAI Bal Acc 75.1%), while other methods showed substantial degradation.

For Alzheimer’s Disease, Sequential Learning with Experience Replay and Diagnosis-pretrained MT again achieved the highest prognosis performance (AUROC 0.851 and 0.848). For diagnosis, Sequential Learning with Experience Replay preserved strong performance (macro AUROC 0.758), whereas Single-cohort MT and Diagnosis-pretrained MT suffered significant degradation (macro AUROC 0.53).

For Breast Cancer, Sequential Learning with Experience Replay and Diagnosis-pretrained MT yielded the best prognosis performance (AUROC 0.876 and 0.878 respectively). Critically, Sequential Learning with Experience Replay maintained diagnostic metrics nearly identical to the single-task reference model (Bal Acc 50.4% vs 50.9%), while all other multitask methods showed poor to degraded diagnostic performance.

The robustness of the Sequential Learning with Experience Replay approach was further confirmed in the subgroup analyses (Fig 3). Across the various Knee OA and Alzheimer’s Disease subgroups, this method consistently achieved prognostic performance that was comparable or superior to the dedicated single-task reference model. In contrast, simpler strategies like ‘Single-cohort MT’ often showed a notable degradation in performance. These results highlight Sequential Learning with Experience Replay as an optimal multitask approach, effectively preventing catastrophic forgetting while achieving top-tier prognostic performance.

**Fig 3.**
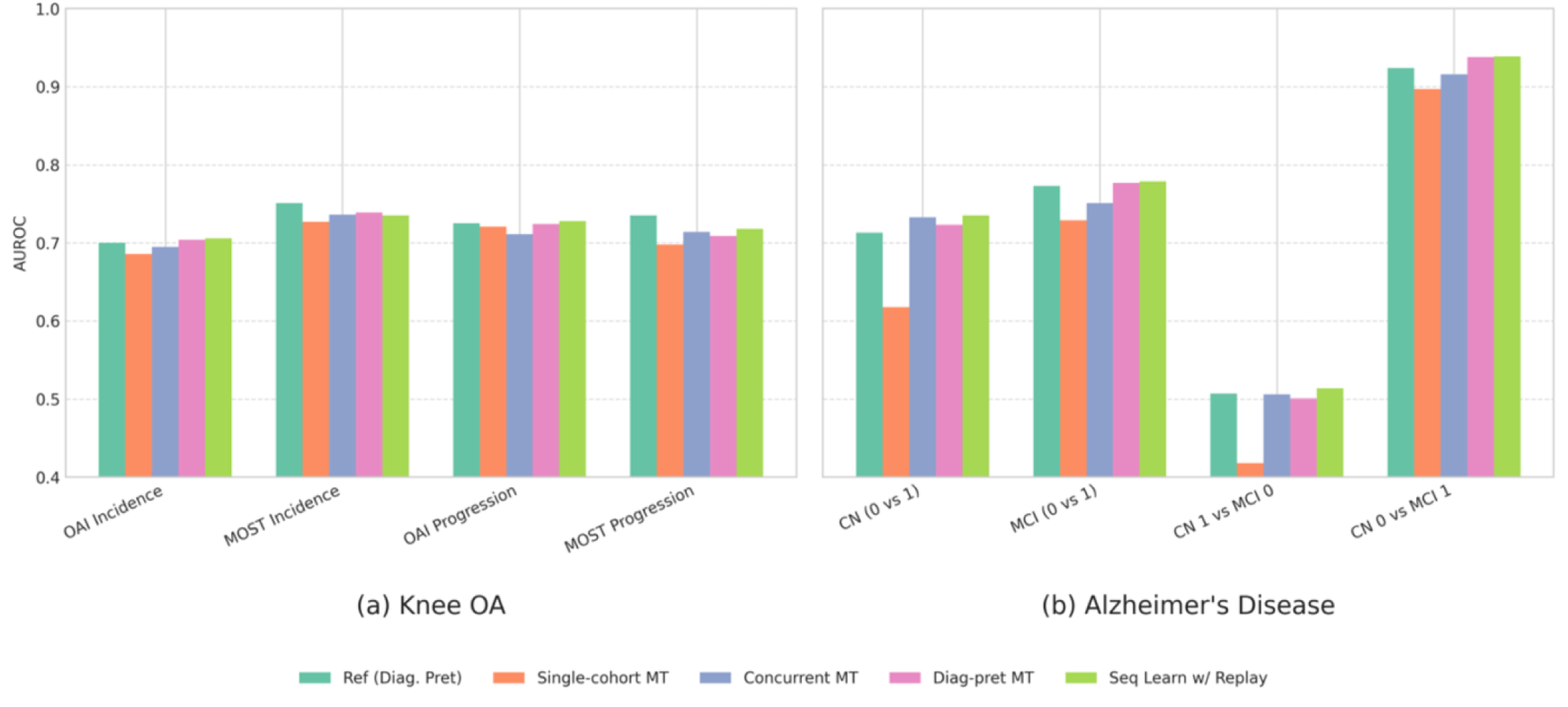
Performance comparison of various multitask learning approaches for prognosis prediction (AUROC) within specific disease subgroups. (a) Knee OA subgroups (OAI/MOST Incidence and Progression) and (b) Alzheimer’s Disease subgroups (ADNI CN/MCI transitions) are shown. Strategies compared include a single-task reference model (’Ref (Diag. Pret)’); Multitask training using only the prognosis cohort (’Single-cohort MT’); Concurrent training on diagnosis and prognosis cohorts (’Concurrent MT’); Multitask training on the prognosis cohort after diagnostic pretraining (’Diag-pret MT’); and Sequential Learning with Experience Replay (’Seq Learn w/ Replay’). Sequential Learning with Replay demonstrates robust performance across most subgroups, often matching or exceeding the dedicated single-task reference model. **Full numerical results, including standard deviations, are available in S7 and S8 Tables.**

## 4 Discussion

Our experiments demonstrate two main findings across the studied conditions: (1) diagnostic pretraining improves prognosis prediction and provides stability, and (2) a sequential learning approach with experience replay performed best among the multitask strategies evaluated.

Diagnostic-pretrained initialization improved prognostic discrimination and generalization to an external cohort and across demographic/clinical subgroups. The advantage remained after controlling for diagnostic severity (Fig. 2), indicating that pretraining helps models capture relevant prognostic features beyond simply using diagnosis severity as a proxy for risk.

Furthermore, our evaluation of joint diagnosis-prognosis approaches showed that sequential learning with experience replay performed best. Across knee OA and AD subgroups, it reached prognostic performance comparable to dedicated, diagnosis-pretrained single-task models, while crucially, also preserving diagnostic performance. This dual success effectively mitigates the critical issue of catastrophic forgetting (Table 3, Fig. 4).

**Fig 4.**
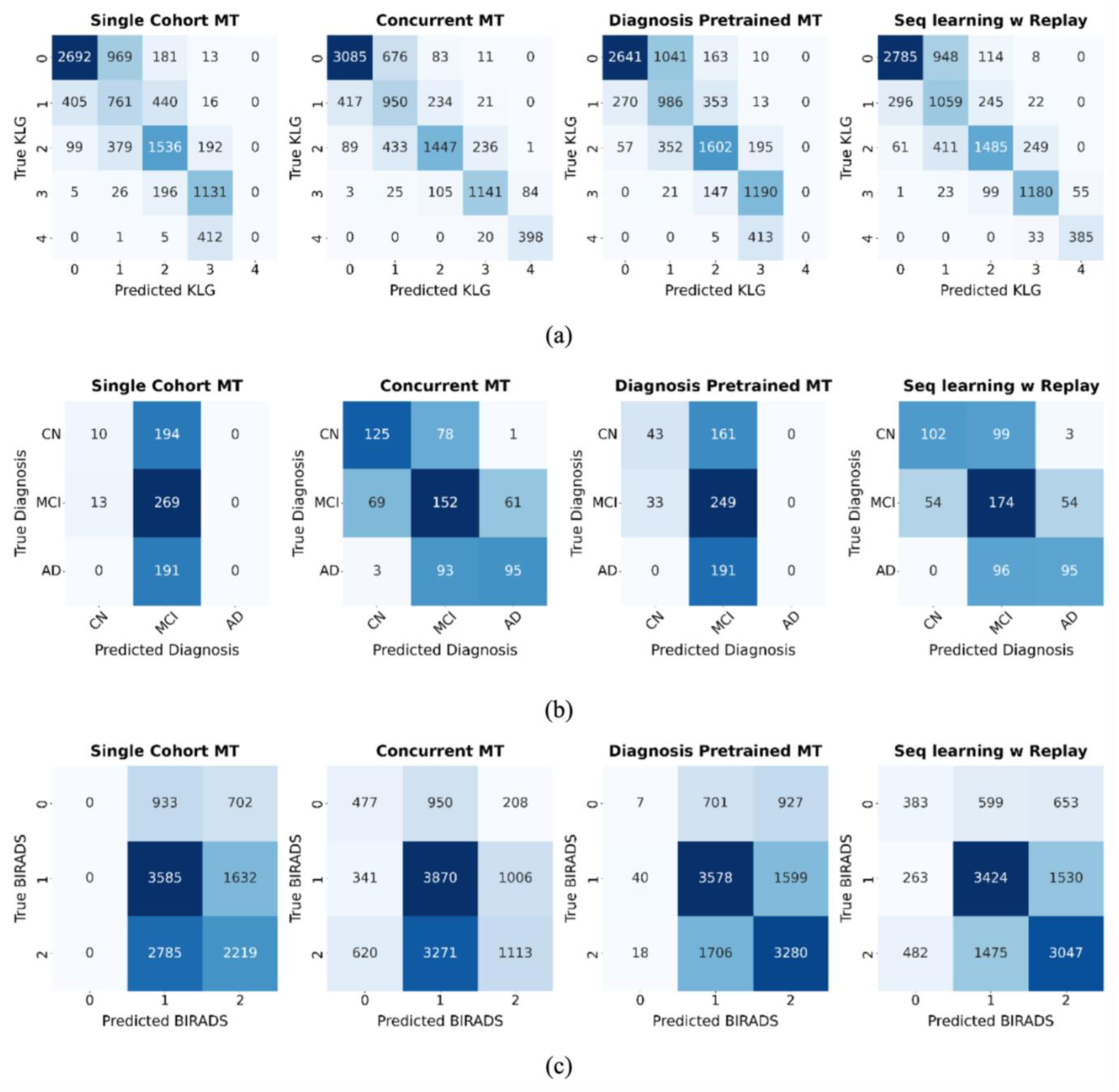
Confusion matrices for the diagnosis task across different multitask learning approaches. (a) Knee OA KLG prediction, (b) Alzheimer’s diagnosis, and (c) Breast Cancer BI-RADS prediction. The matrices show that methods like ‘Diagnosis Pretrained MT’ forget how to classify rare classes (e.g., KLG 4, AD, BI-RADS 0) after prognosis fine-tuning. In contrast, ‘Seq learning w/ Replay’ retains knowledge across all classes, demonstrating its effectiveness at mitigating catastrophic forgetting.

In contrast, the ‘Single-cohort MT’ and ‘Diagnosis-pretrained MT’ variants were evaluated to represent simpler strategies from prior works and serve as a crucial baseline. Their performance starkly illustrates the severity of catastrophic forgetting. Lacking access to the full, unbiased diagnostic dataset during fine-tuning, these approaches suffered a sharp degradation in diagnostic ability. They effectively forgot knowledge from pretraining, including how to classify advanced disease stages in knee OA/AD and the rare class BI-RADS 0 in breast cancer (Fig 4).

This predictable decay in the baseline models highlights the essential value of the replay mechanism. By continually refreshing knowledge from the original task, experience replay preserves diagnostic competence - including for rare classes, while the model learns the new prognosis objective. The consistent success of this strategy across three physiologically distinct conditions (musculoskeletal, neurological, and oncological) suggests its broad applicability for other progressive diseases where longitudinal data is limited.

This work improves upon prior multitask learning strategies by demonstrating that the Single-cohort MT approach, similar to that used by Tiulpin et al. [14] and Leung et al. [16], is suboptimal for knee OA prognosis. Similarly in breast cancer, this study extends the findings of Wu et al. [13], showing that pretraining on diagnosis BI-RADS labels benefits not just breast cancer detection but also long-term prognosis.

Several limitations warrant discussion. First, our knee OA and AD datasets derive from standardized research protocols, potentially lacking the variability of routine clinical practice. Second, the inherent class imbalance in progressive disease studies affects all conditions studied and may impact model performance in deployment. Third, a practical consideration for the sequential learning with experience replay strategy is its reliance on access to the original diagnosis dataset, which may not always be available when models are shared. It is important to note, however, that this limitation applies only to the multitask fine-tuning stage. The initial, performance gains from using a diagnosis-pretrained model for standard fine-tuning - our first key finding remains an applicable benefit of our work, even without access to the original data for replay.

From a clinical perspective, these findings are relevant. A single, unified model that supports both diagnosis and prognosis mirrors routine workflow, assess current status, then plan forward. Such a model can aid counseling by presenting a fuller view - from present severity to future risk and can support personalization. For example, flagging patients at high risk of rapid OA progression for earlier intervention, or using screening mammograms to guide breast-cancer surveillance intensity. Because the sequential learning strategy preserves diagnostic competence, the model remains a dependable diagnostic aid while adding prognostic value, critical for trust and clinical adoption. In addition, producing diagnosis and prognosis together enables cross-verification at the point of care: clinicians can check whether current findings align with predicted risk and more closely review discordant cases, which may support trust and uptake. We view this as a usability/trust benefit rather than a validated safety outcome. Future prospective studies are needed to quantify its impact on decision quality. Additional future work should test these models on larger, more heterogeneous datasets from diverse care settings to assess generalizability beyond the research cohorts used here.

In conclusion, our work demonstrates an effective approach to extracting more value from limited longitudinal datasets by leveraging larger diagnosis datasets for prognostic model training.

## Data Availability

Osteoarthritis Initiative (OAI). OAI data (imaging and clinical) are distributed via the NIMH Data Archive (NDA). Access requires: (i) creating or linking an NDA account (eRA Commons / Login.gov / PIV/CAC) (ii) agreeing to the OAI data access terms within the NDA and (iii) requesting the desired OAI collections through the NDA portal. Detailed instructions are provided on the NDA OAI access pages (https://nda.nih.gov/oai) Multicenter Osteoarthritis Study (MOST). MOST data are available through the NIA Aging Research Biobank. Investigators create a Biobank account and submit a data request in accordance with the Biobank's terms and conditions. (Older UCSF guidance described emailing a signed DUA and request form current distribution is through the Aging Research Biobank.) (https://agingresearchbiobank.nia.nih.gov/) Alzheimer's Disease Neuroimaging Initiative (ADNI). ADNI data are available from the LONI Image & Data Archive upon completion of the web application and acceptance of the ADNI Data Use Agreement. (ADNI: https://adni.loni.usc.edu/data-samples/adni-data/) Breast cancer cohort (NYU Langone Health). This institutional dataset contains protected health information and cannot be posted publicly under the IRB protocol. De-identified data may be shared on reasonable request pending approval by NYU Langone's data governance committee inquiries can be directed to Krzysztof J. Geras. Code. All code for preprocessing and model training will be released in a public repository (license and link to be provided upon acceptance) sufficient to reproduce the analyses reported here.

https://agingresearchbiobank.nia.nih.gov/

https://adni.loni.usc.edu/data-samples/adni-data/

## Acknowledgments

We gratefully acknowledge data provision from the Osteoarthritis Initiative (OAI), an NIH-funded public-private partnership (this manuscript does not necessarily reflect the opinions of OAI investigators or funders); the Multicenter Osteoarthritis Study (MOST), sponsored by the NIH/National Institute on Aging; and the Alzheimer’s Disease Neuroimaging Initiative (ADNI). ADNI is funded by NIH Grant U01 AG024904, DOD ADNI (W81XWH-12-2-0012), and numerous public and private contributions, with a full list of contributors available at adni.loni.usc.edu. The OAI is a public-private partnership comprised of five contracts funded by the National Institutes of Health and private partners including Merck Research Laboratories; Novartis Pharmaceuticals Corporation, GlaxoSmithKline; and Pfizer, Inc. MOST is a nationwide research study sponsored by the National Institutes of Health or National Institute on Aging.

## Supporting Information

**S1 Fig.**
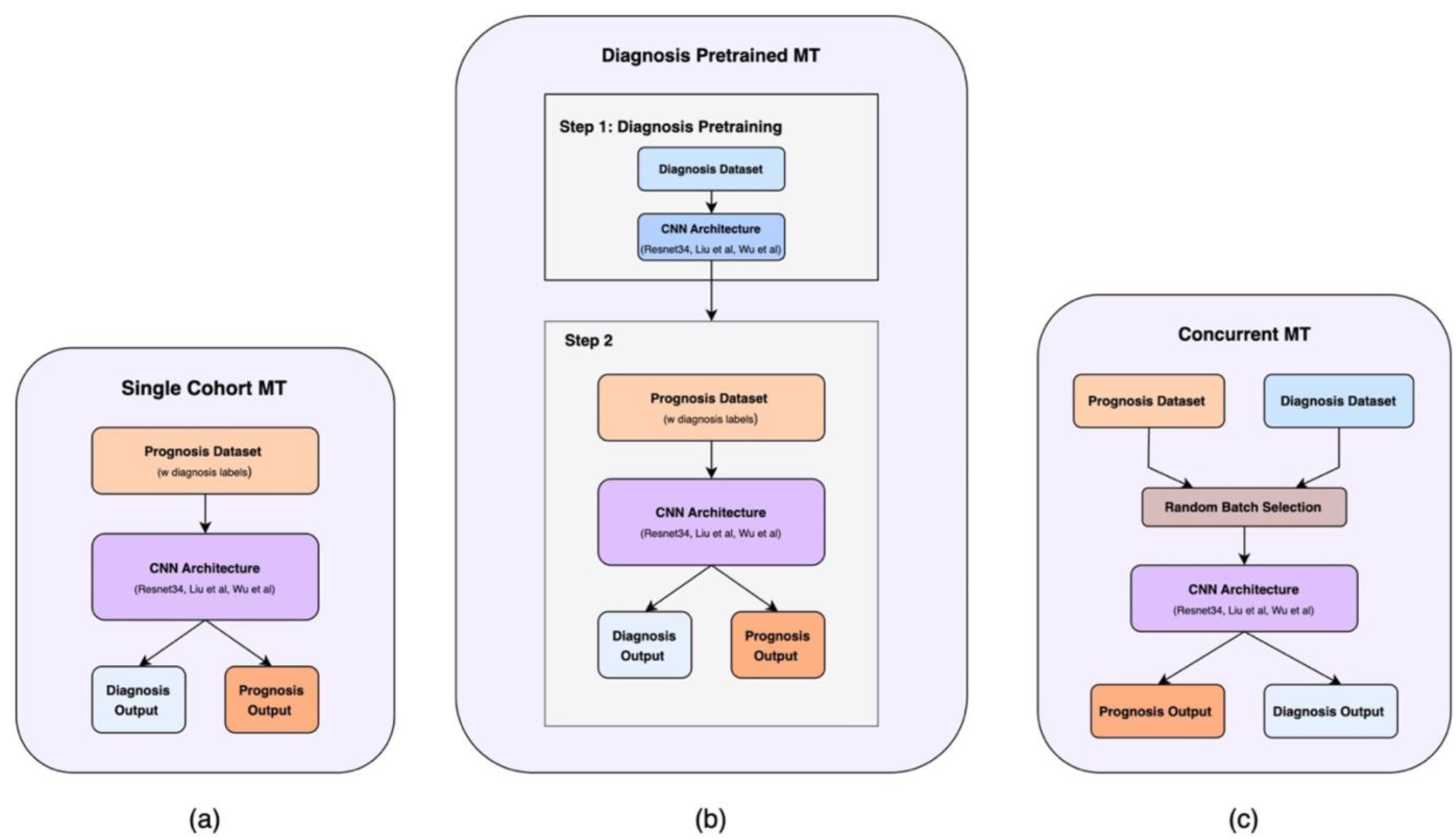
Comparative multitask learning approaches. Visualizations of (a) Single Cohort MT, (b) Diagnosis Pretrained MT, and (c) Concurrent MT.

**S1 Table.** Patient Characteristics in Structural Progression OAI and MOST Cohorts.

| Dataset | Parameters | Men |  | Women |  |
| --- | --- | --- | --- | --- | --- |
|  |  | Patients | Controls | Patients | Controls |
| OAI Dataset | No. of patients | 216 | 707 | 355 | 970 |
|  | No. of scans | 241 | 917 | 421 | 1348 |
|  | Mean age (y) | 62.3±9.0 | 62.1±9.3 | 63.7±8.2 | 62.5±8.9 |
|  | Mean height (m) | 1.8±0.1 | 1.8±0.1 | 1.6±0.1 | 1.6±0.1 |
|  | Mean weight (kg) | 93.1±14.3 | 91.7±14.4 | 80.8±14.8 | 78.2±14.5 |
|  | Mean BMI (kg/m<sup>2</sup>) | 30.1±4.0 | 29.3±4.0 | 30.6±5.2 | 29.6±5.3 |
|  | **Ethnicity** |  |  |  |  |
|  | White | 184 | 595 | 253 | 721 |
|  | Black | 26 | 97 | 90 | 230 |
|  | Asian | 2 | 4 | 3 | 5 |
|  | Other nonwhite | 4 | 11 | 9 | 14 |
| MOST Dataset | No. of patients | 203 | 231 | 359 | 386 |
|  | No. of scans | 231 | 282 | 427 | 495 |
|  | Mean age (y) | 62.8±8.5 | 63.2±8.4 | 64.2±7.6 | 63.5±7.3 |
|  | Mean height (m) | 1.8±0.1 | 1.8±0.1 | 1.6±0.1 | 1.6±0.1 |
|  | Mean weight (kg) | 100.9±19.8 | 98.3±17.8 | 86.3±18.5 | 85.0±18.2 |
|  | Mean BMI (kg/m<sup>2</sup>) | 32.0±5.8 | 31.1±5.4 | 32.7±6.8 | 31.9±6.6 |
|  | **Ethnicity** |  |  |  |  |
|  | White | 177 | 197 | 288 | 302 |
|  | Black | 22 | 32 | 63 | 82 |
|  | Other | 4 | 2 | 8 | 2 |

\*Note: Mean data are presented as mean±standard deviation. BMI = Body Mass Index, MOST= Multi-Center Osteoarthritis Study, OAI= Osteoarthritis Initiative. Number of patients are shown for ethnicity data.

**S2 Table.** Patient Characteristics in Structural Incidence OAI and MOST Cohorts.

| Dataset | Parameters | Men |  | Women |  |
| --- | --- | --- | --- | --- | --- |
|  |  | Patients | Controls | Patients | Controls |
| OAI Dataset | No. of patients | 149 | 1103 | 271 | 1336 |
|  | No. of scans | 160 | 1737 | 296 | 2171 |
|  | Mean age (y) | 61.0±8.7 | 59.6±9.4 | 60.3±8.6 | 60.5±9.0 |
|  | Mean height (m) | 1.8±0.1 | 1.8±0.1 | 1.6±0.1 | 1.6±0.1 |
|  | Mean weight (kg) | 91.0±14.9 | 91.0±14.9 | 77.2±14.2 | 71.3±13.3 |
|  | Mean BMI (kg/m<sup>2</sup>) | 29.1±4.1 | 28.1±3.8 | 29.3±4.8 | 27.1±4.8 |
|  | **Ethnicity** |  |  |  |  |
|  | White | 133 | 975 | 208 | 1121 |
|  | Black | 13 | 110 | 56 | 180 |
|  | Asian | 1 | 4 | 4 | 13 |
|  | Other nonwhite | 2 | 12 | 2 | 21 |
| MOST Dataset | No. of patients | 179 | 600 | 326 | 795 |
|  | No. of scans | 196 | 945 | 371 | 1304 |
|  | Mean age (y) | 61.3±7.8 | 60.6±7.8 | 62.4±8.0 | 61.1±7.7 |
|  | Mean height (m) | 1.8±0.1 | 1.8±0.1 | 1.6±0.1 | 1.6±0.1 |
|  | Mean weight (kg) | 99.8±17.1 | 93.7±15.1 | 82.6±15.6 | 77.4±14.1 |
|  | Mean BMI (kg/m<sup>2</sup>) | 31.3±5.3 | 29.6±4.5 | 30.9±5.9 | 28.8±5.2 |
|  | **Ethnicity** |  |  |  |  |
|  | White | 157 | 573 | 270 | 694 |
|  | Black | 19 | 73 | 50 | 89 |
|  | Other | 3 | 10 | 3 | 12 |

*\*Note: Mean data are presented as mean±standard deviation. BMI = Body Mass Index, MOST=* *Multi-Center Osteoarthritis Study, OAI= Osteoarthritis Initiative. Number of patients are shown* *for ethnicity data.*

**S3 Table.** Demographic Characteristics and Cognitive Status Distribution across the ADNI Prognosis Cohort.

| Parameters | Men |  | Women |  |
| --- | --- | --- | --- | --- |
|  | Patients | Controls | Patients | Controls |
| No. of patients | 89 | 116 | 59 | 101 |
| No. of scans | 204 | 325 | 139 | 246 |
| Age | 75.7 ± 7.3 | 75.5 ± 6.2 | 73.5 ± 7.1 | 75.2 ± 5.9 |
| Cognitive Normal (CN) | 19 | 180 | 12 | 175 |
| Mild Cognitive Impaired (MCI) | 185 | 145 | 127 | 71 |
| **Ethnicity** |  |  |  |  |
| White | 85 | 107 | 55 | 93 |
| Black | 2 | 5 | 3 | 8 |
| Asian | 2 | 4 | 1 | 0 |

*Note: Data shows age (mean ± standard deviation) and number of subjects by cognitive status (CN vs MCI) for both controls (no disease progression, label = 0) and patients (Disease progression, label = 1).*

**S4 Table.** Breast cancer Patient distribution in the Prognosis Cohort.

| Parameters | Patients | Controls |
| --- | --- | --- |
| No. of patients | 2,319 | 2,837 |
| No. of exams | 3,000 | 3,000 |
| Mean age (y) | 61.5±11.4 | 56.4±10.8 |
| **Exam-level BI-RADS** |  |  |
| 0 | 453 | 383 |
| 1 | 962 | 1,411 |
| 2 | 1,599 | 1,189 |
| others | 26 | 17 |
| **Density** |  |  |
| Almost entirely fatty | 112 | 260 |
| Scattered areas of fibroglandular density | 821 | 1,200 |
| Heterogeneously dense | 1,119 | 1,124 |
| Extremely dense | 179 | 221 |
| Unknown | 88 | 32 |
| **Ethnicity** |  |  |
| White | 1,588 | 1,774 |
| Black | 220 | 248 |
| Asian | 115 | 159 |
| Other Nonwhite | 396 | 656 |

**S5 Table.** Comparison of AUROC performance for models trained on progression and incidence, using different initializations.

| Approach | Incidence (KL- 0,1) |  |  |  | Progression (KL - 2,3) |  |  |  |
| --- | --- | --- | --- | --- | --- | --- | --- | --- |
|  | OAI<br>AUROC | OAI<br>AUPRC | MOST<br>AUROC | MOST<br>AUPRC | OAI<br>AUROC | OAI<br>AUPRC | MOST<br>AUROC | MOST<br>AUPRC |
| ImageNet<br>pretrained | 0.697 ±<br>0.02 | 0.222 ±<br>0.017 | 0.707 ±<br>0.019 | 0.380 ±<br>0.023 | 0.726 ±<br>0.011 | 0.561 ±<br>0.016 | 0.699 ±<br>0.011 | 0.672 ±<br>0.009 |
| Diagnosis<br>pretrained | 0.699 ±<br>0.009 | 0.186 ±<br>0.011 | 0.742 ±<br>0.013 | 0.414 ±<br>0.027 | 0.718 ±<br>0.011 | 0.571 ±<br>0.015 | 0.731 ±<br>0.005 | 0.722 ±<br>0.009 |

**S6 Table.** Detailed AUROC analysis comparing model performance across cognitive status subgroups.

| Approach | Within Group |  | Cross Group |  |
| --- | --- | --- | --- | --- |
|  | CN (0 vs 1) | MCI (0 vs 1) | CN 1 vs MCI 0 | CN 0 vs MCI 1 |
| Random initialization | 0.681 ± 0.103 | 0.750 ± 0.028 | 0.461 ± 0.065 | 0.888 ± 0.021 |
| Diagnosis Pretrained | 0.713 ± 0.037 | 0.773 ± 0.016 | 0.507 ± 0.029 | 0.924 ± 0.009 |

**S7 Table.** Detailed metrics for various training strategies for progression and incidence prediction tasks on OAI and MOST Datasets.

| Approach | Incidence (KL-0,1) |  |  |  | Progression (KL-2,3) |  |  |  |
| --- | --- | --- | --- | --- | --- | --- | --- | --- |
|  | OAI | OAI | MOST | MOST | OAI | OAI | MOST | MOST |
|  | AUROC | AUPRC | AUROC | AUPRC | AUROC | AUPRC | AUROC | AUPRC |
| Diagnosis pretrained Ref | 0.700 ±<br>0.017 | 0.220 ±<br>0.019 | 0.751 ±<br>0.006 | 0.433 ±<br>0.008 | 0.725 ±<br>0.003 | 0.567 ±<br>0.008 | 0.735 ±<br>0.002 | 0.731 ±<br>0.003 |
| Single Cohort MT | 0.686 ±<br>0.026 | 0.223 ±<br>0.033 | 0.727 ±<br>0.048 | 0.406 ±<br>0.052 | 0.721 ±<br>0.013 | 0.552 ±<br>0.012 | 0.698 ±<br>0.010 | 0.663 ±<br>0.015 |
| Concurrent MT | 0.695 ±<br>0.021 | 0.242 ±<br>0.026 | 0.736 ±<br>0.011 | 0.405 ±<br>0.017 | 0.711 ±<br>0.014 | 0.532 ±<br>0.010 | 0.714 ±<br>0.013 | 0.702 ±<br>0.009 |
| Diagnosis pretrained MT | 0.704 ±<br>0.009 | 0.233 ±<br>0.016 | 0.739 ±<br>0.011 | 0.422 ±<br>0.018 | 0.724 ±<br>0.006 | 0.566 ±<br>0.008 | 0.709 ±<br>0.010 | 0.715 ±<br>0.007 |
| Seq Learning w Replay | 0.706 ±<br>0.012 | 0.209 ±<br>0.011 | 0.735 ±<br>0.007 | 0.397 ±<br>0.018 | 0.728 ±<br>0.013 | 0.570 ±<br>0.010 | 0.718 ±<br>0.016 | 0.713 ±<br>0.016 |

**S8 Table.** Detailed progression prediction performance (AUROC) across cognitive status subgroups for multitask methods.

| Approach | Within Group |  | Cross Group |  |
| --- | --- | --- | --- | --- |
|  | CN (0 vs 1) | MCI (0 vs 1) | CN 1 vs MCI 0 | CN 0 vs MCI 1 |
| Diagnosis pretrained Ref | 0.713 ± 0.037 | 0.773 ± 0.016 | 0.507 ± 0.029 | 0.924 ± 0.009 |
| Single cohort MT | 0.618 ± 0.053 | 0.729 ± 0.020 | 0.418 ± 0.029 | 0.897 ± 0.024 |
| Concurrent MT | 0.733 ± 0.042 | 0.751 ± 0.014 | 0.506 ± 0.028 | 0.916 ± 0.021 |
| Diagnosis pretrained MT | 0.723 ± 0.038 | 0.777 ± 0.013 | 0.501 ± 0.030 | 0.938 ± 0.009 |
| Seq learning w replay | 0.735 ± 0.040 | 0.779 ± 0.007 | 0.514 ± 0.022 | 0.939 ± 0.012 |

## References

1. Cross M, Smith E, Hoy D, Nolte S, Ackerman I, Fransen M, Bridgett L, Williams S, Guillemin F, Hill CL, Laslett LL. The global burden of hip and knee osteoarthritis: estimates from the global burden of disease 2010 study. Annals of the rheumatic diseases. 2014 Jul 1;73(7):1323–30.

2. Steinmetz JD, Culbreth GT, Haile LM, Rafferty Q, Lo J, Fukutaki KG, Cruz JA, Smith AE, Vollset SE, Brooks PM, Cross M. Global, regional, and national burden of osteoarthritis, 1990–2020 and projections to 2050: a systematic analysis for the Global Burden of Disease Study 2021. The Lancet Rheumatology. 2023 Sep 1;5(9):e508–22.

3. Rasmussen J, Langerman H. Alzheimer’s disease–why we need early diagnosis. Degenerative neurological and neuromuscular disease. 2019 Dec 24:123–30.

4. Arnold M, Morgan E, Rumgay H, Mafra A, Singh D, Laversanne M, Vignat J, Gralow JR, Cardoso F, Siesling S, Soerjomataram I. Current and future burden of breast cancer: Global statistics for 2020 and 2040. The Breast. 2022 Dec 1;66:15–23.

5. Allemani C, Matsuda T, Di Carlo V, Harewood R, Matz M, Nikšić M, Bonaventure A, Valkov M, Johnson CJ, Estève J, Ogunbiyi OJ. Global surveillance of trends in cancer survival 2000–14 (CONCORD-3): analysis of individual records for 37 513 025 patients diagnosed with one of 18 cancers from 322 population-based registries in 71 countries. The Lancet. 2018 Mar 17;391(10125):1023–75.

6. LeCun Y, Bottou L, Bengio Y, Haffner P. Gradient-based learning applied to document recognition. Proceedings of the IEEE. 2002 Aug 6;86(11):2278–324.

7. Tiulpin A, Thevenot J, Rahtu E, Lehenkari P, Saarakkala S. Automatic knee osteoarthritis diagnosis from plain radiographs: a deep learning-based approach. Scientific reports. 2018 Jan 29;8(1):1727.

8. Liu S, Masurkar AV, Rusinek H, Chen J, Zhang B, Zhu W, Fernandez-Granda C, Razavian N. Generalizable deep learning model for early Alzheimer’s disease detection from structural MRIs. Scientific reports. 2022 Oct 17;12(1):17106.

9. Liu S, Yadav C, Fernandez-Granda C, Razavian N. On the design of convolutional neural networks for automatic detection of Alzheimer’s disease. In Machine learning for health workshop 2020 Apr 30 (pp. 184–201). PMLR.

10. Castellano G, Esposito A, Lella E, Montanaro G, Vessio G. Automated detection of Alzheimer’s disease: A multi-modal approach with 3D MRI and amyloid PET. Scientific Reports. 2024 Mar 3;14(1):5210.

11. Suk HI, Lee SW, Shen D, Alzheimer’s Disease Neuroimaging Initiative. Hierarchical feature representation and multimodal fusion with deep learning for AD/MCI diagnosis. NeuroImage. 2014 Nov 1;101:569–82.

12. Geras KJ, Wolfson S, Shen Y, Wu N, Kim S, Kim E, Heacock L, Parikh U, Moy L, Cho K. High-resolution breast cancer screening with multi-view deep convolutional neural networks. arXiv preprint arXiv:1703.07047. 2017 Mar 21.

13. Wu N, Phang J, Park J, Shen Y, Huang Z, Zorin M, Jastrzębski S, Févry T, Katsnelson J, Kim E, Wolfson S. Deep neural networks improve radiologists’ performance in breast cancer screening. IEEE transactions on medical imaging. 2019 Oct 7;39(4):1184–94.

14. Leung K, Zhang B, Tan J, Shen Y, Geras KJ, Babb JS, Cho K, Chang G, Deniz CM. Prediction of total knee replacement and diagnosis of osteoarthritis by using deep learning on knee radiographs: data from the osteoarthritis initiative. Radiology. 2020 Sep;296(3):584–93.

15. Tolpadi AA, Lee JJ, Pedoia V, Majumdar S. Deep learning predicts total knee replacement from magnetic resonance images. Scientific reports. 2020 Apr 14;10(1):6371.

16. Tiulpin A, Klein S, Bierma-Zeinstra SM, Thevenot J, Rahtu E, Meurs JV, Oei EH, Saarakkala S. Multimodal machine learning-based knee osteoarthritis progression prediction from plain radiographs and clinical data. Scientific reports. 2019 Dec 27;9(1):20038.

17. Schiratti JB, Dubois R, Herent P, Cahané D, Dachary J, Clozel T, Wainrib G, Keime-Guibert F, Lalande A, Pueyo M, Guillier R. A deep learning method for predicting knee osteoarthritis radiographic progression from MRI. Arthritis Research & Therapy. 2021 Oct 18;23(1):262.

18. Rajamohan HR, Wang T, Leung K, Chang G, Cho K, Kijowski R, Deniz CM. Prediction of total knee replacement using deep learning analysis of knee MRI. Scientific reports. 2023 Apr 28;13(1):6922.

19. Lee G, Nho K, Kang B, Sohn KA, Kim D. Predicting Alzheimer’s disease progression using multi-modal deep learning approach. Scientific reports. 2019 Feb 13;9(1):1952.

20. Spasov S, Passamonti L, Duggento A, Lio P, Toschi N, Alzheimer’s Disease Neuroimaging Initiative. A parameter-efficient deep learning approach to predict conversion from mild cognitive impairment to Alzheimer’s disease. Neuroimage. 2019 Apr 1;189:276–87.

21. Sun D, Wang M, Li A. A multimodal deep neural network for human breast cancer prognosis prediction by integrating multi-dimensional data. IEEE/ACM transactions on computational biology and bioinformatics. 2018 Feb 15;16(3):841–50.

22. Huang Z, Zhang X, Ju Y, Zhang G, Chang W, Song H, Gao Y. Explainable breast cancer molecular expression prediction using multi-task deep-learning based on 3D whole breast ultrasound. Insights into Imaging. 2024 Sep 19;15(1):227.

23. Nevitt M, Felson D, Lester G. The osteoarthritis initiative. Protocol for the cohort study. 2006 Jun;1(2).

24. Segal NA, Nevitt MC, Gross KD, Hietpas J, Glass NA, Lewis CE, Torner JC. The Multicenter Osteoarthritis Study (MOST): opportunities for rehabilitation research. PM & R: the journal of injury, function, and rehabilitation. 2013 Aug;5(8):10–16.

25. Weiner MW, Kanoria S, Miller MJ, Aisen PS, Beckett LA, Conti C, Diaz A, Flenniken D, Green RC, Harvey DJ, Jack Jr CR. Overview of Alzheimer’s Disease Neuroimaging Initiative and future clinical trials. Alzheimer’s & Dementia. 2025 Jan;21(1):e14321.

26. Kellgren JH, Lawrence J. Radiological assessment of osteo-arthrosis. Ann Rheum Dis. 1957 Dec 1;16(4):494–502.

27. Lee S, Kwon Y, Lee N, Bae KJ, Park S, Kim YH, Cho KH. The prevalence of osteoarthritis and risk factors in the Korean population: The Sixth Korea National Health and Nutrition Examination Survey (VI-1, 2013). Korean Journal of Family Medicine. 2018 Nov 8;40(3):171.

28. Wu N, Phang J, Park J, Shen Y, Kim SG, Heacock L, Moy L, Cho K, Geras KJ. The NYU breast cancer screening dataset v1. 0. New York Univ., New York, NY, USA, Tech. Rep. 2019.

29. Robins A. Catastrophic forgetting, rehearsal and pseudorehearsal. Connection Science. 1995 Jun 1;7(2):123–46.

30. Chaudhry A, Rohrbach M, Elhoseiny M, Ajanthan T, Dokania PK, Torr PH, Ranzato MA. On tiny episodic memories in continual learning. arXiv preprint arXiv:1902.10486. 2019 Feb 27.

31. He K, Zhang X, Ren S, Sun J. Deep residual learning for image recognition. In Proceedings of the IEEE conference on computer vision and pattern recognition 2016 (pp. 770–778).

32. Woo S, Park J, Lee JY, Kweon IS. Cbam: Convolutional block attention module. In Proceedings of the European conference on computer vision (ECCV) 2018 (pp. 3–19).

33. Kingma DP. Adam: A method for stochastic optimization. arXiv preprint arXiv:1412.6980. 2014.

34. Youden WJ. Index for rating diagnostic tests. Cancer. 1950;3(1):32–5.

35. DeLong ER, DeLong DM, Clarke-Pearson DL. Comparing the areas under two or more correlated receiver operating characteristic curves: a nonparametric approach. Biometrics. 1988 Sep 1:837–45.

